# Therapeutic effects of human amnion-derived mesenchymal stem cells on hypercoagulability in a uremic calciphylaxis patient

**DOI:** 10.1101/2023.01.06.22283161

**Authors:** Anning Bian, Xiaoxue Ye, Jing Wang, Ming Zeng, Jiayin Liu, Kang Liu, Song Ning, Yugui Cui, Shaowen Tang, Xueqiang Xu, Yanggang Yuan, Zhonglan Su, Yan Lu, Jing Zhou, Xiang Ma, Guang Yang, Yaoyu Huang, Xiaolin Lv, Ling Wang, Jing Zhao, Xiuqin Wang, Ningxia Liang, Changying Xing, Lianju Qin, Ningning Wang

## Abstract

Calciphylaxis is a rare cutaneous vascular disease with clinical manifestations of intolerable pains, non-healing skin wounds, histologically characterized by calcification, fibrointimal hyperplasia, and thrombosis in microvessels. Currently, there are no approved guidelines for this disease. High prevalence of thrombophilias and hypercoagulable conditions in calciphylaxis patients have been recognized in recent years. Here, we report a case of uremic calciphylaxis patient whom was refractory to conventional treatments and then received a salvage strategy *via* human amnion-derived mesenchymal stem cell (hAMSC) intravenous combined with local application. In order to investigate the therapeutic mechanism of hMASCs from the novel perspective of hypercoagulability, coagulation-related indicators, wound status and quality of life were followed up. Improvement of hypercoagulable condition involving correction of platelet, D-dimer and plasminogen levels, skin regeneration and pain alleviation were revealed after hAMSC administration for one year. We propose that hypercoagulability is the therapeutic target of calciphylaxis patients which can be improved by hAMSC treatment.

## Background

Uremic calciphylaxis, also known as calcific uremic arteriolopathy (CUA), is a rare but life-threating cutaneous vascular disease characterized by rapidly progressing, intensely painful, ischemic skin lesions that occur predominately in patients with end-stage kidney disease(ESKD)^1^. The pathohistologic features include calcification, microthrombosis, fibrointimal hyperplasia of small dermal and subcutaneous arteries and arterioles^2^. An estimated incidence of calciphylaxis in patients undergoing dialysis is reported to range from 0.04% to 4%^3^, with 1-year mortality up to 45%-80%^4, 5^. The sepsis secondary to infected ulcerations is considered the most common cause of death^1,2^. Skin biopsy remains the gold standard for the diagnosis of clinically suspected calciphylaxis.

The pathogenesis of this devastating disease remains uncertain despite the gradually increasing reports. A multidisciplinary approach including wound care, analgesia, anti-inflammatory treatment, elimination of risk factors might be effective, however, non-standard treatment has been established^3, 6^. Due to the stimulation of cell proliferation, inhibition of vascular calcification, neovascularization, anti-inflammation and antioxidative effects, mesenchymal stem cells (MSCs) treatment is considered promising for skin regeneration and kidney repair^7-9^.

Although microvascular calcification is regarded as the basic histopathological characteristic, it is noteworthy that hypercoagulable states seem to be crucial in the development of CUA^6, 10^, and anticoagulants including tissue plasminogen activator or low molecular weight heparin have been reported effective^6, 10-12^. We have reported an innovative therapeutic strategy for a CUA patient *via* intravenous combined with local treatment of hAMSCs, with speculated mechanisms including the inhibition of vascular calcification, the stimulation of neovascularization and myogenesis, as well as anti-inflammation, immunoregulatory actions, and re-epithelialization^13^. The aim of this study is to investigate the effects of hAMSCs on the improvement of hypercoagulation in CUA patient and analyze the possible mechanisms.

## Method

### Case summary

We reported a young female calciphylaxis patient in her 30s underwent peritoneal dialysis (PD), who was admitted to our hospital with multiple skin lesions accompanied by intolerable pain for more than 1 month. The skin of her back, thighs, lower limbs, and buttocks were presented with induration, plaques, purpura, livedo reticularis, and ecchymosis, gradually progressing into malodorous necrotic ulcerations surrounded by leather-like skin. Based on the clinical manifestation, medical history and skin histopathology, the patient was diagnosed as CUA. The comorbidities included secondary hyperparathyroidism (SHPT), skin and soft tissue infections, PD-related tunnel infection, malnutrition and hypertension.

### Salvage treatment strategy with hAMSCs for CUA patient

Due to the ineffectiveness to multiple conventional therapies, she was treated with hAMSCs, a rescue therapeutic strategy approved by the Ethics Committee of The First Affiliated Hospital of Nanjing Medical University, Jiangsu Province Hospital (2018-QT-001). hAMSCs were provided by the State Key Laboratory of Reproductive Medicine in our Hospital^14^. A series of hAMSC preclinical tests including validity and safety assessments were performed before clinical administration^13^. The CUA patient was administered hAMSC intravenously at a dosage of 1.0×10^6^ cells/kg, along with a local intramuscular injection at the edge of the wound (2.0×10^4^ cells/cm^2^) and external application of cell culture supernatants on the wound surface^13^. Detailed flow chart of hAMSC treatment was demonstrated in Figure 1A. Written informed consent was signed prior to hAMSC therapy.

**Figure 1.**
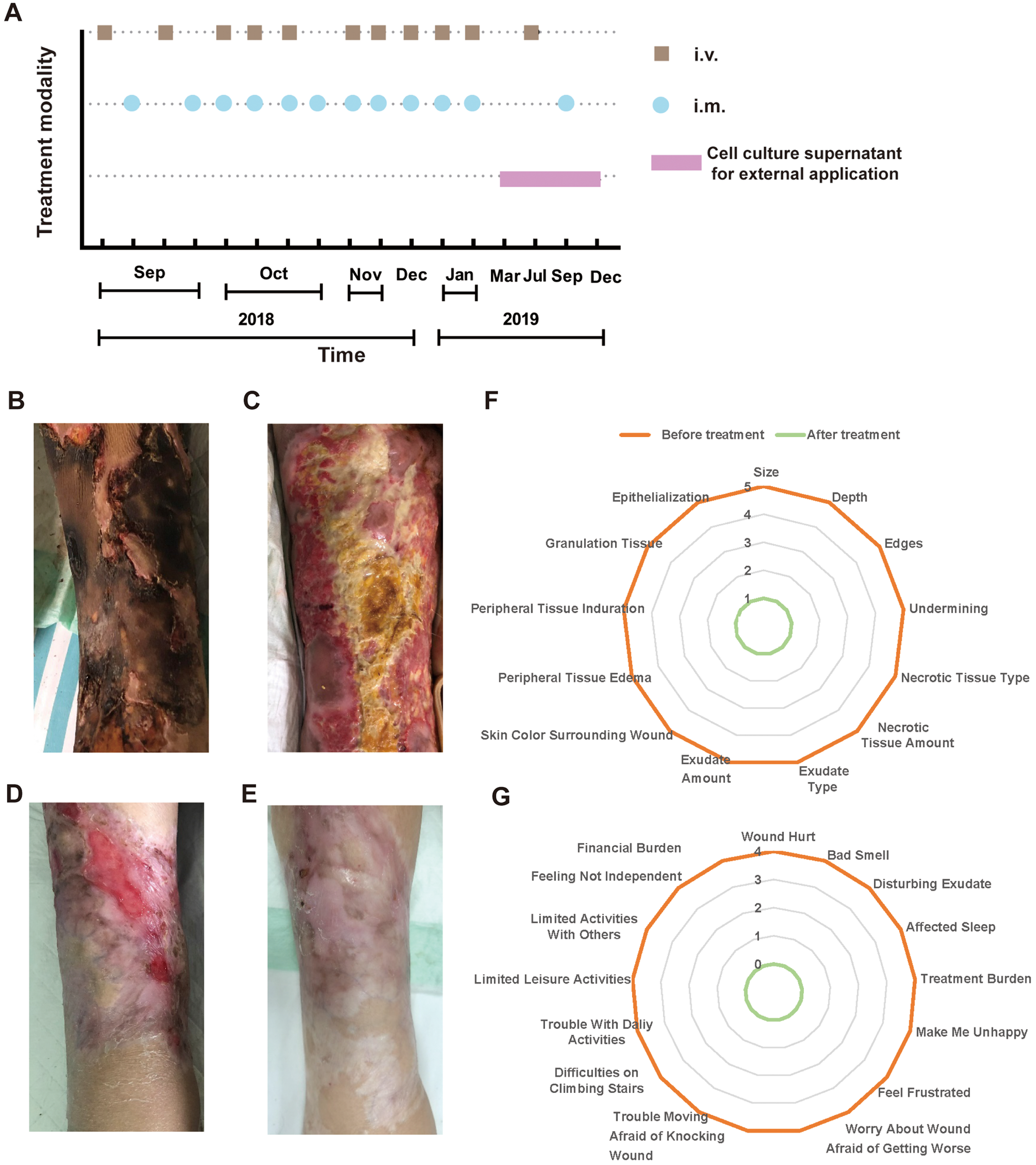
Flow chart of hAMSC treatment and skin regeneration, quality of life in the CUA patient. (A) Flow chart of hAMSC treatment. (B-E) Healing process of the skin lesions on the right thigh during the course of hAMSC treatment. (B) The anterior of right thigh presenting with large areas of irregular black scab, superficial ulceration and slight exudation before hAMSC treatment. (C) Patchy shallow ulcer covered with granulation tissues, pale yellow necrotic tissues and crusting in the center after hAMSC treatment for 2 months and debridement. (D) Striated scar with reddish dry ulcer in the center, pigment reduction in the peripheral, dilated blood vessels and a small amount of local crusting after hAMSC treatment for 9 months. (E) Fully regenerated skin accompanied with irregularly shaped scar, localized hypopigmentation and few scabs in the right thigh after hAMSC treatment for 20 months. (F-G) Wound assessment based on BWAT(F) and quality of life evaluated by Wound-QoL(G) after hAMSC treatment for 15 months. Abbreviations: hAMSC = human amnion-derived mesenchymal stem cell; CUA = calcific uremic arteriolopathy; BWAT = Bates-Jensen Wound Assessment; Wound-QoL = Wound-Quality of Life.

### Observed efficacy indicators

During the course of hAMSC treatment, the dynamic changes of blood parameters including coagulation indicators (platelet, D-dimer, fibrinogen), C reactive protein (CRP) and albumin, clinical symptoms, pain level, and quality of life were followed up. Pain Visual Analog Scale (VAS) was applied to rate pain level on a scale of zero to 10^15^. Bates-Jensen Wound Assessment (BWAT), containing 13 evaluation items, was used to assess wound status during interventions. Each item ranges from 1 to 5 and a total score of 13 represents the healthiest and 65 indicates the unhealthiest characteristic of wound condition^16^. Wound-Quality of Life (Wound-QoL) was implemented to measure quality of life with 17 assessment items divided into three subscales on everyday life, body and psyche. Each item is scored from 0 to 4, representing low to high level of quality of life. Overall score ranges from 0 equal to “not at all” representing the best imaginable to 68 equal to “very much” representing the worst imaginable health status^17^. Detailed scoring items were shown in Figure 1F-G.

## Results

We performed skin biopsy on this CUA patient for the definitive diagnosis. The skin histology revealed focal defect in the epidermis, chronic inflammatory cells infiltration in the dermis, subcutaneous calcification of the mesothelium of small arteries, subintimal fibrosis, thickening of the vessel wall with luminal narrowing and fibrous tissue hyperplasia with hyaline degeneration, in accordance with the pathological manifestations of calciphylaxis. Multidisciplinary therapies involving analgesics, antibiotic administration, wound care, nutrition support, continuous kidney replacement therapy (CKRT), sodium thiosulfate and anticoagulation^1, 2^ were applied, but without improvement in skin lesions and pain relief. Due to lacking of alternative therapeutic options, we rescue this CUA patient with hAMSCs as a candidate^13^.

The skin manifestations, wound status and quality of life were shown in Figure 1. We followed up the healing process of skin lesions on the right thigh before treatment (Figure 1B) and after 2 months (Figure 1C), 9 months (Figure 1D), 20 months (Figure 1E) of hAMSC application. It was noteworthy that the right thigh obtained full skin regeneration when administered hAMSC for 20 months ultimately. Hence, the patient got great relief of wound pain and the scores of pain VAS decreased from 10 to 0 after 15 months hAMSC treatment. Likewise, the score of BWAT decreased from 65 to 13 points, accompanying with substantial improvement in quality of life after hAMSC application for 15 months (Figure 1F-G). Either inflammation status or immune disorder was ameliorated after hAMSC administration for one year, with tumor markers in normal ranges^13^.

As hypercoagulability is a potential risk factor for CUA, we monitored coagulation-related indicators including platelet, D-dimer and fibrinogen (Figure2), which declined immediately after one week hAMSC administration. After hAMSC treatment for 3 months, the levels of platelet and fibrinogen tended to be in normal ranges. After 6 months of hAMSC therapy, D-dimer showed a fluctuating downward trend to physiological level. The inflammation related marker, CRP, decreased substantially after hAMSC therapy for 3 months.

**Figure 2.**
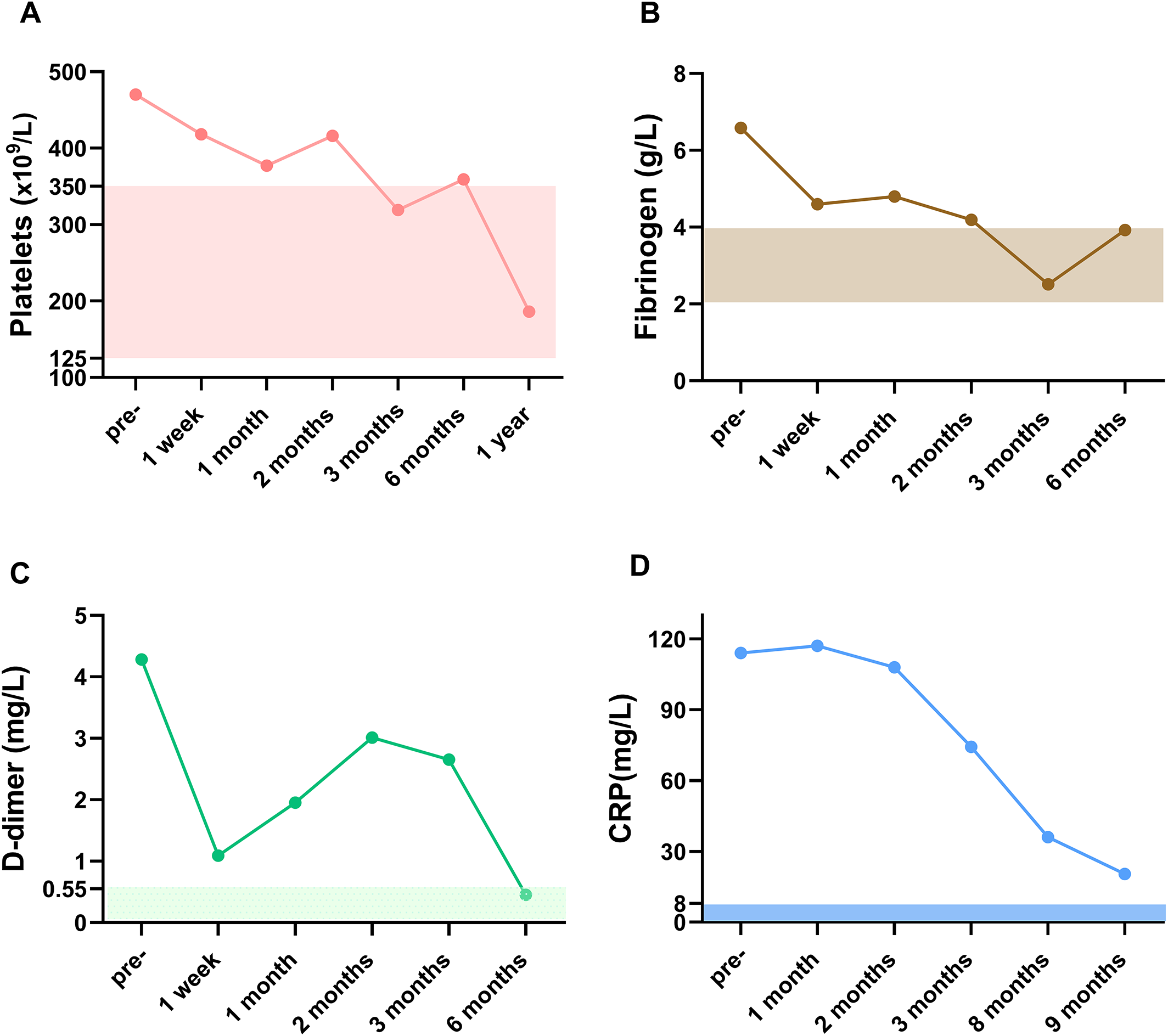
The levels of blood coagulation-related markers of CUA patient during the course of hAMSC treatment. (A)Platelets, (B) Fibrinogen, (C) D-dimer and (D) CRP were followed up. The rectangular shaded area represented the normal reference range for each index and the recommended ranges were based on the normal reference values of laboratory tests. Abbreviations: CUA = calcific uremic arteriolopathy; hAMSC = human amnion-derived mesenchymal stem cell; CRP = C reactive protein

## Dicussion

Calciphylaxis is a rare and severely morbid disorder with high mortality that predominantly affects ESKD patients. Various comorbidities are considered to be the risk factors, involving obesity, female, disordered calcium-phosphate metabolism, hypoalbuminemia, warfarin, thrombophilia associated with protein C and protein S deficiencies^18^. Although clinical and histological descriptions of CUA have been characterized, its pathogenic mechanisms remain poorly understood and the important role of hypercoagulability has been noticed^10, 19^.

There are case reports of calciphylaxis with both hereditary and acquired thrombophilic conditions such as protein S deficiency, protein C deficiency, lupus anticoagulant and antithrombin deficiency^19, 20^. It has been reported that 60% calciphylaxis patients have severe thrombophilia^20^. Harris revealed that 38% and 43% of calciphylaxis patients had decreased protein C and protein S levels, respectively^19^. Additionally, elevated fibrinogen (47%) and D-dimer (41%) levels in the plasma were observed in calciphylaxis cohort^20^. Calciphylaxis patients treated with tissue plasminogen activator had approximately 30% greater survival than controls, but significant difference was not observed^11^. Therapeutic success in a case of calciphylaxis is thought to be associated with low molecular weight heparin for hypercoagulable condition^12^. Hence, as a potential risk factor, hypercoagulability plays an important role in the pathogenic mechanism and is proposed to be the treatment target for calciphylaxis.

D-dimer is a plasmin-derived soluble degradation product of cross-linked fibrin, served as a valuable marker of coagulation and fibrinolysis activation. Of note, D-dimer is a sensitive marker for detection of thrombosis, but its specificity is not ideal^21^. Fibrinogen, the soluble precursor of fibrin, is a major determinate of blood viscosity and red blood cells aggregation. Elevated fibrinogen levels may provide a hypercoagulable state through increasing the aggregation and reactivity of platelet^22^. In this case, the plasma D-dimer and fibrinogen levels were both elevated before hAMSC treatment, which represented an underlying hypercoagulable condition. The patient presented a high inflammatory status before hAMSC treatment, as revealed by the substantially elevated CRP and leukocytes levels. Increasing evidence points out that systemic inflammation will induce the activation of coagulation system, which may result from tissue factor (TF)-mediated thrombin generation or the dysfunction of anticoagulant pathway, such as the impaired protein C system^23^. Compared with physiological serum level of albumin, platelet aggregation was significantly higher in the presence of low serum albumin level^24^. This case had hypoalbuminaemia at the onset of hAMSC administration. Thus, either acquired thrombophilic conditions like protein C and protein S deficiency or hypercoagulability induced by inflammation and hypoalbuminaemia may be correlated with the development of CUA.

In this case after hAMSC treatment, the levels of plasma D-dimer and fibrinogen decreased immediately after one week administration and gradually showed a tendency to normal range accompanied by controlled inflammatory state. We speculated that hAMSCs may exert therapeutic roles in CUA by correcting hypercoagulable conditions. Netsch et al^25^demonstrated that MSCs originating from bone marrow, adipose and cord blood inhibited the agonist-induced activation and aggregation of platelets in platelet-rich plasma (PRP) and whole blood, expressing anticoagulant properties. The underlying mechanism was responsible for the CD73-mediated adenosine generation by MSCs^25^. Additionally, placenta MSCs were reported to act a protective role in agonist-induced platelet activation, inhibiting thrombosis and atherosclerosis, partially through decreasing CD36-mediated platelet activation^26^. In this case, we presume that hAMSCs may alleviate hypercoagulability of CUA either directly or indirectly through improving the inflammatory state and hypoalbuminemia. Of note, the thrombogenic risk induced by MSCs associated with expression of procoagulant molecules such as TF has drawn attention in recent studies^27^. In our preclinical acute toxicity animal tests, high-dosage hAMSCs can cause pulmonary embolism and death in mice. During hAMSC treatment for this CUA patient, the intravenous dosage(1.0×10^6^ cells/kg) was lower than the maximum tolerated dosage in C57BL/6 mice (equivalent to 6.0 × 10^6^ cells/kg in humans) and in Sprague–Dawley rats (equivalent to 20.0 × 10^6^ cells/kg in humans)^13^. Hence, the intravenous dose we currently administered is considered safe and will not directly result in the complication of embolism.

### Limitations

This is a case study of hAMSC treatment playing crucial role in improving hypercoagulability in CUA patient, thus the causal relationships between hypercoagulability and calciphylaxis can not be concluded in this report. A complete thrombophilia tests involving protein C, protein S, antithrombin III or other related hypercoagulability indicators were not performed in this case.

## Conclusion

Our findings implied that skin lesions and inflammatory status were improved *via* salvage hAMSC therapy based on the conventional multidisciplinary therapies, which may partly due to the improvement of hypercoagulability in the calciphylaxis patient. There is strong support that identification of coagulation profile and a thorough search for both congenital and acquired thrombophilias should be assessed for the new onset calciphylaxis patients. In order to improve our understanding of this complex disease, future research should focus on early causative factors for hypercoagulability in CUA and the identification of driving forces for the progress of vasculopathy as dichotomic event. Prospective multi-institutional collaborative efforts to clarify the safety and efficacy of novel anticoagulants and hAMSC therapy in calciphylaxis patients are suggested when there was risk of thrombosis predisposing to ulceration.

## Data Availability

All data produced in the present study are available upon reasonable request to the authors.

## Abbreviations

hAMSC: human amnion-derived mesenchymal stem cell
CUA: calcific uremic arteriolopathy
ESKD: end-stage kidney disease
PD: peritoneal dialysis
SHPT: secondary hyperparathyroidism
VAS: Visual Analog Scale
BWAT: Bates-Jensen Wound Assessment
Wound-QoL: Wound-Quality of Life
CKRT: continuous kidney replacement therapy
CRP: C reactive protein
TF: tissue factor
PRP: platelet-rich plasma.

## Acknowledgements

The authors thank the patient, her family and all medical staff who participated in the treatment and data collection. The study was supported by the International Society of Nephrology (ISN) Mentorship Program and the authors thank Professor Marcello Tonelli (University of Calgary, Canada) for his helpful comments on the draft of the manuscript.

## Authors’ contributions

WNN, QLJ, and LJY conceived and directed the project, designed the hAMSC clinical treatment, and support the research funding. XCY provided clinical guidance. WXQ and LNX provided research ethical issues and administrative guidance. TSW provided guidance for clinical research. QLJ, NS, CYG and Jing Zhou prepared hAMSC cell line, performed quality control. MX donated human amniotic membrane tissues. WNN, WJ, ZM, LK, XXQ, YYG, SZL, LY, YG, HYY, LXL, WL and Jing Zhao performed clinical management and multidisciplinary team rescue of the patient. BAN, YXX, WJ, QLJ and WNN analyzed the data, wrote, revised and submitted the manuscript.

## Availability of data and materials

The data are available from the corresponding author on reasonable request.

## Ethics approval and consent to participate

The study was performed after an approval by the Ethics Committee of The First Affiliated Hospital of Nanjing Medical University, Jiangsu Province Hospital (2018-QT-001). Healthy pregnant women in their 20s or 30s who provided written informed consent donated human amniotic membranes, which was approved by the Ethics Committee of Jiangsu Province Hospital (2012-SR-128).

## Consent for publication

The patient and her family gave the consent for publication of the data obtained in the present study.

